# Association of Mortality and Aspirin Prescription for COVID-19 Patients at the Veterans Health Administration

**DOI:** 10.1101/2020.12.13.20248147

**Authors:** Thomas F Osborne, Zachary P Veigulis, David M Arreola, Satish Mahajan, Eliane Röösli, Catherine M Curtin

## Abstract

There is growing evidence that thrombotic and inflammatory pathways contribute to the severity of COVID-19. Common medications such as aspirin, that mitigate these pathways, may decrease COVID-19 mortality. This assessment was designed to quantify the correlation between aspirin and mortality for COVID-19 positive patients in our care. Data from the Veterans Health Administration national electronic health record database was utilized for the evaluation. Veterans from across the country with a first positive COVID-19 polymerase chain reaction lab result were included in the evaluation which comprised 28,350 patients from March 2, 2020 to September 13, 2020 for the 14-day mortality cohort and 26,346 patients from March 2, 2020 to August 28, 2020 for the 30-day mortality cohort. Patients were matched via propensity scores and the odds of mortality were then compared. Among COVID-19 positive Veterans, preexisting aspirin prescription was associated with a statistically and clinically significant decrease in overall mortality at 14-days (OR 0.38, 95% CI 0.32-0.46) and at 30-days (OR 0.38, 95% CI 0.33-0.45), cutting the odds of mortality by more than half. Findings demonstrated that pre-diagnosis aspirin prescription was strongly associated with decreased mortality rates for Veterans diagnosed with COVID-19. Prospective evaluation is required to more completely assess this correlation and its implications for patient care.

## Introduction

Veterans Health Administration (VA) is the largest integrated healthcare system in the United States (U.S.) and enrolled Veterans represent a population at increased risk of poor COVID-19 outcomes due to older age and multiple comorbidities ^1-5^. As part of our continual quality improvement efforts, we have been developing and validating predictive models to optimize care strategies. Our recent work demonstrated that existing electronic health record (EHR) data can be utilized at VA to assess risk in the form of the Care Assessment Need (CAN) score for different groups of Veterans including those battling COVID-19 ^6-7^. In the process of optimizing COVID-19 specific predictive models, we identified a strong correlation between preexisting aspirin prescription and decreased all-cause mortality. Given the urgency of this crisis, and the potential for drug repurposing to improve outcomes, this analysis is our in-depth evaluation of the correlation between aspirin and mortality for COVID-19 positive Veterans.

## Materials and methods

### Design

Data was obtained from the VA national Corporate Data Warehouse (CDW), a central relational database repository that aggregates EHR records from 1,255 VA health care facilities across the U.S. This database includes enrolled Veterans as well as a very small number of non-Veterans such as qualified spouses. Our work to refine VA risk assessment tools for COVID-19 outcomes started with binary logistic regression modeling of mortality on common variables available in our EHR such as comorbidities and pre-diagnosis medications. For appropriate variable size, a medication class or a specific medication was included in the assessment if utilized by more than 10% of the cohort population. This resulted in the inclusion of 31 medication variables, of which 17 were specific medications and 14 were separate medication classes. These medication variables are listed in the supplemental material (S1). There was a strong correlation between pre-diagnosis use of aspirin and decreased all-cause mortality in each of the logistic regression models we evaluated. We utilized log odds of at least −0.05 in mortality as an inclusion criterion for medications in the assessment, and of the 31 medications evaluated, pre-diagnosis aspirin prescription was the only one that met this criterion (mortality decreased by 36% with a log odds of −0.45). Retrospective analysis was then performed on both aspirin and non-aspirin groups for 14-day and 30-day mortality. The CAN score was utilized to compare baseline characteristics of the different cohorts.

### Patients

CDW lab tables were queried to identify the first positive COVID-19 polymerase chain reaction results for patients. For this evaluation, the time of the first positive COVID-19 test administration was considered the time of diagnosis. Test results performed outside of VA were not included in the evaluation to reduce potential bias of different risk patients receiving care outside VA, test variability, and incomplete access to outside records. Patients under the age of 18 and over the age of 105 were excluded from the assessment, which represented two patients. Patients without a calculated CAN score within 6 months prior to their first positive COVID-19 lab results were also excluded since a patient’s CAN score was used to inform health status and patients not actively utilizing VA care will not have a CAN score calculated, which represented 7,020 patients. The final analysis included 28,350 patients with a first positive COVID-19 polymerase chain reaction lab result from March 2, 2020 to September 13, 2020 for the 14-day mortality cohort, and 26,346 patients from March 2, 2020 to August 28, 2020 for the 30-day mortality cohort. Each cohort has a different end date to allow for appropriate follow-up time window to elapse before analysis could be performed for all-cause mortality.

### Variables

The aspirin cohort was defined as those patients who had an active aspirin prescription and medication delivery by the pharmacy at the time of a positive COVID-19 lab test. If a patient had no refills at the time of diagnosis, then the prescription was only considered active if it was filled up to 30 days prior to the positive COVID-19 lab test. Patients who had non-VA aspirin prescriptions documented as active medications in the EHR were also considered active and were included in the treatment group. All other COVID-19 patients, with no active aspirin prescription documented, were assigned to the control cohort. This methodology was adopted based on previous work utilizing VA EHR data to assess aspirin use ^8^.

Patients’ health risk was controlled for by using the VA CAN score. The CAN score is a tool that assesses patients’ risk of morbidity and mortality using a wide array of data available in the EHR, including socio-demographics, clinical diagnoses, vital signs, medications, lab values, and health care utilization data. Our previous work has established that the CAN score successfully risk-stratifies COVID-19 patient outcomes with the strongest performance demonstrated in predicting mortality ^6-7^. The CAN score ranges from 0 to 99, with a higher score representing a greater risk patient. We utilized the CAN 1-year mortality model (version 2.5), which is automatically computed weekly based on all living Veterans who actively receive outpatient care services from VA.

The primary outcomes were 14-day and 30-day all-cause mortality within or outside of hospital care. Mortality was identified by date and time of death listed in CDW. The mortality time windows for the evaluation began at the date of the first VA positive COVID-19 test.

### Statistical Analysis

For both the 14-day and 30-day mortality outcomes that were evaluated, unadjusted odds ratios using contingency tables were first computed to quantify the difference in mortality between the aspirin treatment and control group. Given the retrospective observational nature of the data, leading to significant demographic differences in the treatment and control groups, propensity score matching was then applied to reduce confounding effects. To do so, the treatment and control group were matched one-to-one on the unscaled covariates of age, gender, and CAN (1-year mortality) with the RStudio “MatchIt” library (V3.6.2) using the commonly used caliper setting of 0.1. Race (White vs. other) was not included as a covariate in the matching algorithm since our assessment demonstrated that it was not significantly associated with the treatment assignment nor the outcome. New contingency tables and adjusted odds ratios were then computed on the matched cohort for both the all-cause 14-day and 30-day mortality outcomes. Fixing the control cohort as the reference group, odds ratios are reported for the aspirin treatment cohort as point estimates with 95% confidence intervals.

This quality improvement project received a Determination of Non-Research from Stanford IRB (Stanford University, Stanford, CA, USA), as well as by VA determination.

## Results

### Patient characteristics

For the 14-day observational window, a total of 28,350 patients were identified with a positive COVID-19 test at VA from March 2, 2020 to September 13, 2020 that met our inclusion criteria. For the 30-day observational window, a total of 26,346 patients were identified with a positive COVID-19 test at VA from March 2, 2020 to August 28, 2020. In both the cohorts, approximately 24% were prescribed aspirin prior to COVID-19 diagnosis. The majority of both the unmatched cohorts are male, which is expected for our VA population. Both cohorts were virtually the same age (58 years each) and in the same health as represented by the CAN score (CAN of 44 each) and Charlson Comorbidity Index (CCI of 2.7 each). More than half of all patients had hypertension, more than 30% had chronic pulmonary disease and diabetes, and 15% had congestive heart failure (Table 1).

**Table 1.** Characteristics of the two patient cohorts.

|  | **14-Day Mortality**<br>**n = 28350** | **30-Day Mortality**<br>**n = 26346** |
| --- | --- | --- |
|  | **Mean (SD)** | **Mean (SD)** |
| **Age Years** | 58.44 (16.68) | 58.41 (16.70) |
| **Charlson Comorbidity Index<sup>8</sup>** | 2.66 (3.12) | 2.66 (3.13) |
| **Care Assessment Need (CAN) 1 Year Mort.** | 44.43 (31.85) | 44.45 (31.89) |
|  | **n (%)** | **n (%)** |
| **Male** | 25276 (89.2) | 23487 (89.1) |
| **Hypertension** | 17561 (61.9) | 16308 (61.9) |
| **Chronic Pulmonary Disease** | 9149 (32.3) | 8480 (32.2) |
| **Congestive Heart Failure** | 4113 (14.5) | 3851 (14.6) |
| **Diabetes** | 10353 (36.5) | 9619 (36.5) |
| **American Indian or Alaska Native** | 339 (1.2) | 302 (1.2) |
| **Asian** | 337 (1.2) | 312 (1.2) |
| **Black or African American** | 9883 (35.5) | 9374 (36.2) |
| **Native American or other Pacific Islander** | 300 (1.1) | 276 (1.1) |
| **Not reported** | 2254 (8.0) | 2085 (7.9) |
| **White** | 15237 (53.7) | 13997 (53.1) |

### Treatment and control groups

For the treatment and control groups, the differences in age, gender, and CAN were resolved after applying propensity score matching (Table 2, 3). The scatter plots of the propensity score as a function of the covariates age, gender, and CAN demonstrate very closely fitting curves to further confirm this assessment (Figure S2).

**Table 2.**
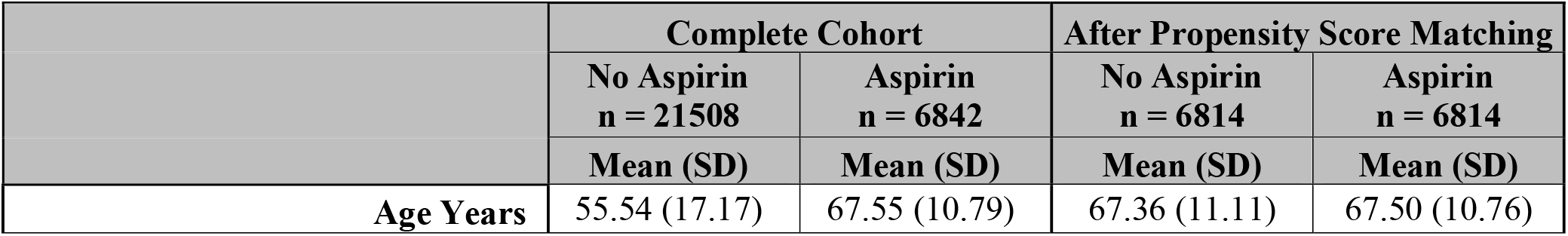
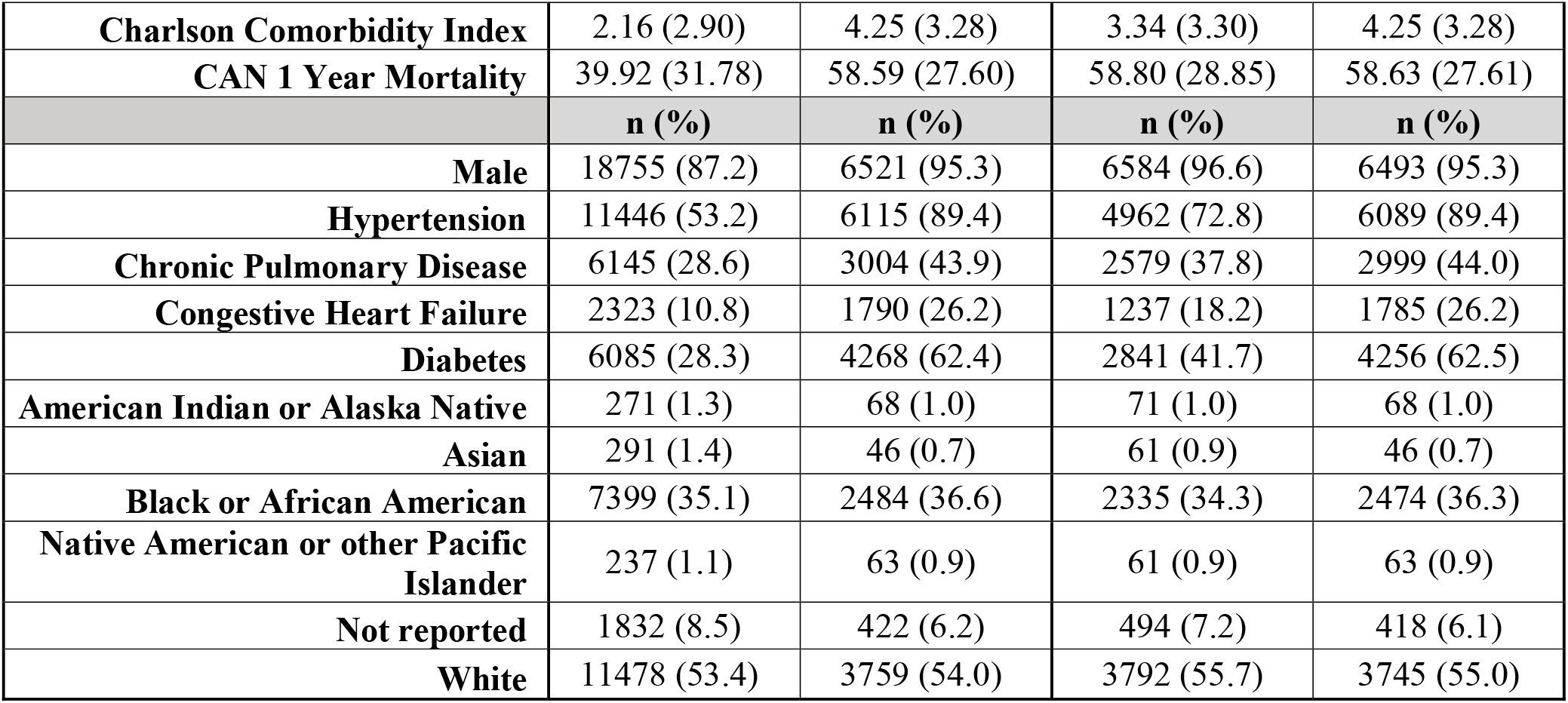
Patient characteristics for the 14-day mortality cohort before and after propensity score matching.

|  | **Complete Cohort** |  | **After Propensity Score Matching** |  |
| --- | --- | --- | --- | --- |
|  | **No Aspirin**<br>**n = 21508**<br>**Mean (SD)** | **Aspirin**<br>**n = 6842**<br>**Mean (SD)** | **No Aspirin**<br>**n = 6814**<br>**Mean (SD)** | **Aspirin**<br>**n = 6814**<br>**Mean (SD)** |
| **Age Years** | 55.54 (17.17) | 67.55 (10.79) | 67.36 (11.11) | 67.50 (10.76) |
| **Charlson Comorbidity Index** | 2.16 (2.90) | 4.25 (3.28) | 3.34 (3.30) | 4.25 (3.28) |
| **CAN 1 Year Mortality** | 39.92 (31.78) | 58.59 (27.60) | 58.80 (28.85) | 58.63 (27.61) |
|  | **n (%)** | **n (%)** | **n (%)** | **n (%)** |
| **Male** | 18755 (87.2) | 6521 (95.3) | 6584 (96.6) | 6493 (95.3) |
| **Hypertension** | 11446 (53.2) | 6115 (89.4) | 4962 (72.8) | 6089 (89.4) |
| **Chronic Pulmonary Disease** | 6145 (28.6) | 3004 (43.9) | 2579 (37.8) | 2999 (44.0) |
| **Congestive Heart Failure** | 2323 (10.8) | 1790 (26.2) | 1237 (18.2) | 1785 (26.2) |
| **Diabetes** | 6085 (28.3) | 4268 (62.4) | 2841 (41.7) | 4256 (62.5) |
| **American Indian or Alaska Native** | 271 (1.3) | 68 (1.0) | 71 (1.0) | 68 (1.0) |
| **Asian** | 291 (1.4) | 46 (0.7) | 61 (0.9) | 46 (0.7) |
| **Black or African American** | 7399 (35.1) | 2484 (36.6) | 2335 (34.3) | 2474 (36.3) |
| **Native American or other Pacific Islander** | 237 (1.1) | 63 (0.9) | 61 (0.9) | 63 (0.9) |
| **Not reported** | 1832 (8.5) | 422 (6.2) | 494 (7.2) | 418 (6.1) |
| **White** | 11478 (53.4) | 3759 (54.0) | 3792 (55.7) | 3745 (55.0) |

**Table 3.**
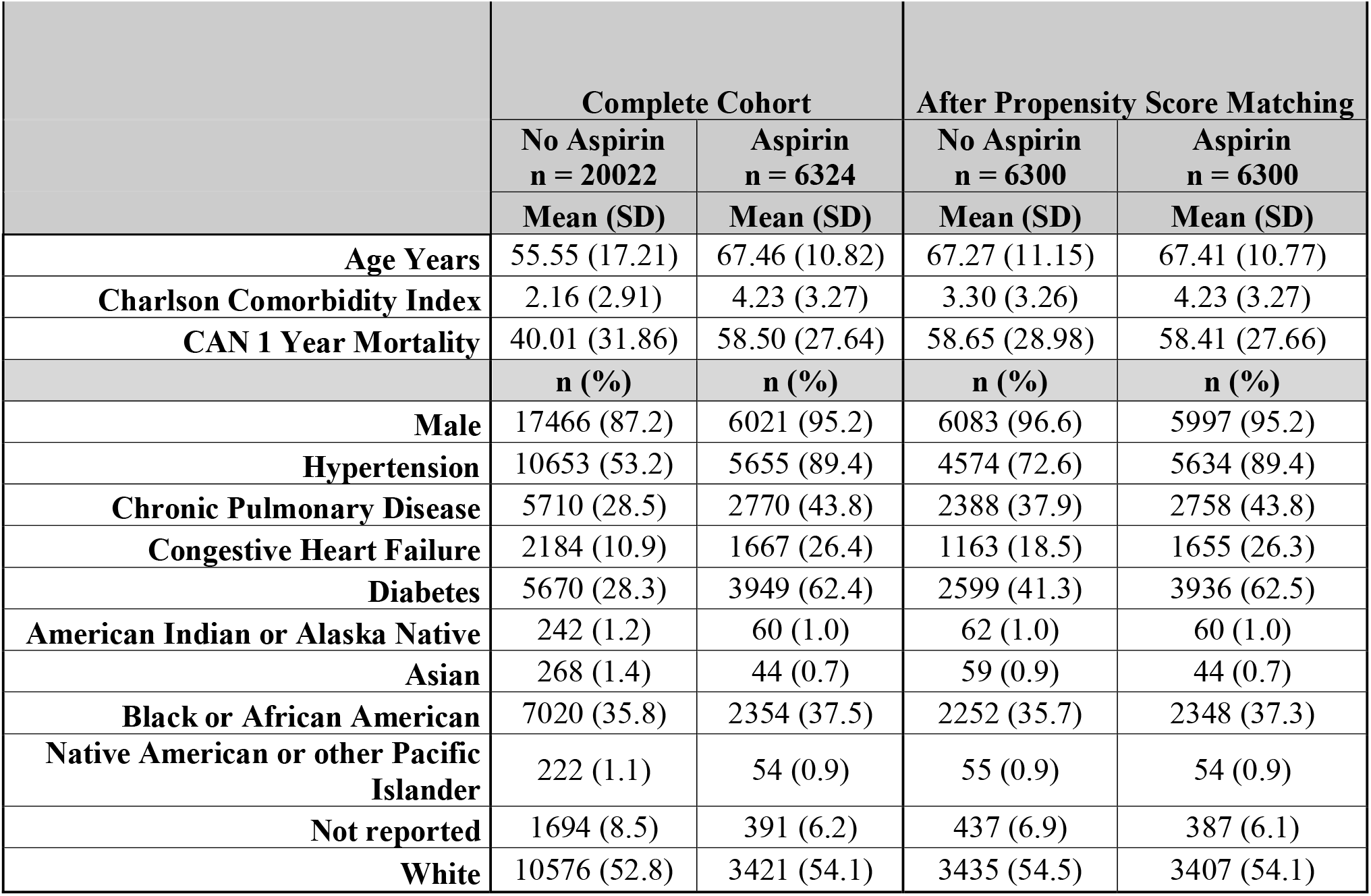
Patient characteristics for the 30-day mortality cohort before and after propensity score matching.

|  | **Complete Cohort** |  | **After Propensity Score Matching** |  |
| --- | --- | --- | --- | --- |
|  | **No Aspirin**<br>**n = 20022** | **Aspirin**<br>**n = 6324** | **No Aspirin**<br>**n = 6300** | **Aspirin**<br>**n = 6300** |
|  | **Mean (SD)** | **Mean (SD)** | **Mean (SD)** | **Mean (SD)** |
| **Age Years** | 55.55 (17.21) | 67.46 (10.82) | 67.27 (11.15) | 67.41 (10.77) |
| **Charlson Comorbidity Index** | 2.16 (2.91) | 4.23 (3.27) | 3.30 (3.26) | 4.23 (3.27) |
| **CAN 1 Year Mortality** | 40.01 (31.86) | 58.50 (27.64) | 58.65 (28.98) | 58.41 (27.66) |
|  | **n (%)** | **n (%)** | **n (%)** | **n (%)** |
| **Male** | 17466 (87.2) | 6021 (95.2) | 6083 (96.6) | 5997 (95.2) |
| **Hypertension** | 10653 (53.2) | 5655 (89.4) | 4574 (72.6) | 5634 (89.4) |
| **Chronic Pulmonary Disease** | 5710 (28.5) | 2770 (43.8) | 2388 (37.9) | 2758 (43.8) |
| **Congestive Heart Failure** | 2184 (10.9) | 1667 (26.4) | 1163 (18.5) | 1655 (26.3) |
| **Diabetes** | 5670 (28.3) | 3949 (62.4) | 2599 (41.3) | 3936 (62.5) |
| **American Indian or Alaska Native** | 242 (1.2) | 60 (1.0) | 62 (1.0) | 60 (1.0) |
| **Asian** | 268 (1.4) | 44 (0.7) | 59 (0.9) | 44 (0.7) |
| **Black or African American** | 7020 (35.8) | 2354 (37.5) | 2252 (35.7) | 2348 (37.3) |
| **Native American or other Pacific Islander** | 222 (1.1) | 54 (0.9) | 55 (0.9) | 54 (0.9) |
| **Not reported** | 1694 (8.5) | 391 (6.2) | 437 (6.9) | 387 (6.1) |
| **White** | 10576 (52.8) | 3421 (54.1) | 3435 (54.5) | 3407 (54.1) |

### Mortality

COVID-19 positive patients with active aspirin prescriptions have a significantly decreased risk of mortality as indicated by unadjusted odds ratios of 0.68 (95% CI of 0.57-0.80) at 14 days, and 0.68 (95% CI of 0.59-0.77) at 30 days after diagnosis. Accounting for age, gender, and CAN score via propensity score matching, the adjusted odds of dying while having an aspirin prescription was 0.38 (95% CI of 0.32-0.46) at 14-days, and 0.38 (95% CI of 0.33-0.45) at 30-days (Table 4). In the propensity-matched cohort, these adjusted odds ratios represent a decrease in 14-day mortality from 6.3% (control group) to 2.5% (treatment group) and a decrease in 30-day mortality from 10.5% (control group) to 4.3% (treatment group).

**Table 4.** Unadjusted and adjusted odds ratios for the 14-day and 30-day mortality aspirin cohorts (95% confidence interval). All odds ratio p-values are < 0.001.

|  | 14-Day Mortality | 30-Day Mortality |
| --- | --- | --- |
| **Unadjusted OR** | 0.68 (0.57-0.80) | 0.68 (0.59-0.77) |
| **Adjusted OR** | 0.38 (0.32-0.46) | 0.38 (0.33-0.45) |

## Discussion

As part of our COVID-19 quality improvement efforts at VA, we have been developing and validating COVID-19 prediction models to optimize Veteran care. In the process of evaluating logistic regression models, we discovered a strong correlation between preexisting aspirin prescription and decreased mortality in COVID-19 positive Veterans. This observation also confirms a recent study of 400 patients, which found aspirin use to be associated with lower COVID-19 mortality ^9^. This exciting finding in our large national cohort presents the possibility that aspirin, a well-known and inexpensive medication, could be an important complementary tool in the COVID-19 medical armamentarium.

The impact of propensity score matching on the odds ratio points to a strong confounding of aspirin’s treatment effect by the covariates age, gender, and CAN score. Accounting for these risk factors, that are commonly associated with aspirin prescriptions, findings demonstrated that baseline aspirin prescriptions imparted a strong decrease in mortality, essentially cutting the risk of an adverse outcome by more than half in Veterans. In addition, these results are statistically and clinically significant for mortality outcomes measured in both the 14-day and 30-day timeframes.

The associated benefit of aspirin on Veterans in this disease may be related to several factors. A leading possibility is that aspirin’s systemic antithrombotic effects could disrupt the increasingly recognized life-threatening risk of thrombotic events related to COVID-19. As an example, a recent study of autopsy findings in twelve COVID-19 patients found 58% had deep vein thrombosis and four had pulmonary embolus as the direct cause of death ^10^. There is also an increased rate of acute ischemic strokes in COVID-19 patients, which is felt to be a consequence of the hypercoagulable state associated with the infection ^11, 12^. Notably, this evaluation presents positive associations for a population of patients who were on aspirin at the time of diagnosis with COVID-19, and not as a new treatment after becoming critically ill. This may be a crucial distinction because the aspirin platelet inhibition mechanism is beneficial to prevent clot formation but not thrombolysis of an existing clot ^13^. Therefore, if COVID-19 hypercoagulability induced thrombotic events, such as myocardial infarction, pulmonary embolism, and stroke, are an important cause of acute decompensation, the presence of the antiplatelet effects of aspirin before COVID-19 infection could be protective. Likewise, the anti-inflammatory effect of aspirin may also have an independent or synergistic beneficial effect. There are other potential pathways to consider in which aspirin could positively impact patients with COVID-19 such as inhibiting virus replication ^14^.

There are cautions to be considered when discussing aspirin use in the setting of COVID-19. Early in this pandemic, there was concern that NSAIDs could be related to adverse cardiovascular and pulmonary outcomes and there was an editorial published that suggested potential harm with NSAID prescription for COVID-19 patients ^15^. However, the World Health Organization subsequently published a brief report stating that: “…there is no evidence of severe adverse events, acute health care utilization, long-term survival, or quality of life in patients with COVID-19, as a result of the use of NSAID” ^16^. Nevertheless, there are many aspirin contraindications to take into consideration such as in the setting of bleeding disorders, as well as disseminated intravascular coagulation, which can result in uncontrolled bleeding, and in children because of the risk of Reye’s syndrome.

There are important strengths of this assessment. Importantly, our data source included a large number of Veterans, spanning a diverse geographic range, from an integrated longitudinal EHR database. In addition, because VA practices purposeful adverse patient selection, many Veterans rely on VA for OTC medications. Therefore, our database offers a broader scope of care and a more complete record of outpatient aspirin use, which is a crucial variable. However, there are also Veterans with better financial means, who may not rely on VA for OTC medications, and it is possible that some of these patients may not have reported their aspirin use to their clinicians even though medication reconciliation is a national VA system-wide policy for clinical visits and the VA EHR database is designed to include non-VA prescribed OTC medications. Since those with better financial means tend to have better healthcare outcomes, this could result in an underestimation of the beneficial association of aspirin ^17,18^. The relative recent emergence of this pandemic highlights an important limitation of all COVID-19 assessments such as reporting of mortality occurring outside of VA facilities which may be delayed and could also underestimate the results.

There are several areas that deserve additional assessment as more data becomes available. For example, sub-cohort analysis of the relative impact of different dosages of aspirin, as well as the effect of less common anticoagulation medications, will become more statistically significant over time as datasets increase in size. Importantly, as a retrospective evaluation, we cannot establish direct cause and effect, only correlation that deserves dedicated controlled trial to assess the potential of aspirin as a drug repurposing option for this population.

## Conclusion

Aspirin prescription was discovered to be strongly associated with decreased mortality rates for COVID-19 positive patients enrolled at VA. Additional prospective evaluation is required to more completely understand this correlation and the potential implications for improved care.

## Data Availability

Data cannot be shared publicly because it involves sensitive human subject information. Data may be available for researchers who meet the criteria for access to confidential data after evaluation from affiliated IRB and VA Research and Development Committees. As a VA national legal policy (VHA Directive1605.01), VA will only share patient data if there is a fully executed contractual agreement in place for the specific project. A common contractual mechanism utilized for this type of sharing is a Cooperative Research and Development (CRADA) agreement. These contracts are typically negotiated in collaboration with VA national Office of General Council (OGC) and attorneys from the collaborating institution. These national sharing policies and standards also apply to deidentified data. In addition, if a contract is in place allowing sharing of deidentified data outside of VA, then VA national policy (VHA Directive 1605.01), states that deidentification certification needs to be met by Expert Determination. The expert determination requires independent assessment from an experienced master or PhD in biostatistics, from a third party not involved in the project, and may require outside funding to support. In addition, for an outside entity to perform research on VA patient data, IRB as well as VA Research and Development Committee approval is required for the specific project. Data requests may be sent to: VA Information Resource Center (VIReC) Building 18 Hines VA Hospital (151V) 5000 S. 5th Avenue Hines, IL 60141-3030 708-202-2413 708-202- 2415 (fax).

## Acknowledgements

We are grateful for the consultative insight of Reese H. Clark, MD and Todd Wagner, PhD.

---

**S1 Table.** The 31 medication variables utilized in the logistic regression analysis. If a unique medication was present in more than 10% of the cohort, it was included as its own variable. All other medications were grouped as “other” by drug class. This resulted in the inclusion of 31 variables across 14 separate medication classes.

|  |
| --- |
| Atorvastatin |
| Other_AntiLipemic |
| Acetaminophen |
| Aspirin |
| Other_NO_Angalgesic |
| Ibuprofen |
| Naproxen |
| Meloxicam |
| Other_NSAIDs |
| AntiDep |
| Metformin |
| Other_HypoGlycemic |
| Insulin |
| Metoprolol |
| Other_Beta |
| Amlodipine |
| Other_CalcBlock |
| Lisinopril |
| Other_AceInhib |
| Losartan |
| Other_AngioInhib |
| BronchDilat |
| Omeprazole |
| Pantoprazole |
| Other_Gastric |
| Tamsulosin |
| Other_AlphaBlock |
| Gabapentin |
| Other_AntiConvul |
| Opioid |
| Glucocorticoids |

**S2 Figure.**
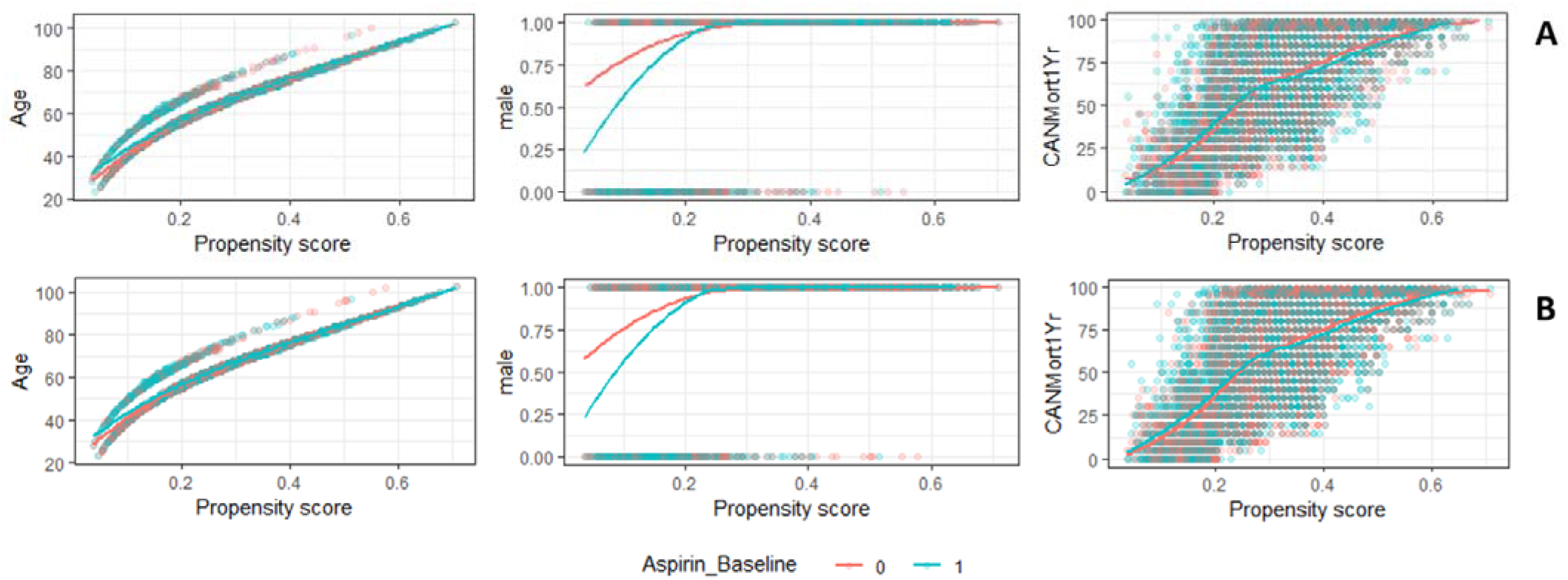
Scatter plots with fitted lines of the association between the propensity score and the matched covariates (age, gender and CAN 1-year mortality score) for the 30-day mortality cohort (A) and the 14-day mortality cohort (B).

## References

1. Kizer KW, Demakis JG, Feussner JR. Reinventing VA health care: systematizing quality improvement and quality innovation. Medical care 2000:I7–I16.

2. Wang J, Cidade M, Larsen M, et al. 2018 survey of veteran enrollees’ health and use of health care. Department of Veterans Affairs Rockville, MD; 2019.

3. About VHA. Washington, DC: Veterans Health Administration, July 2019. (https://www.va.gov/health/aboutvha.asp.). Accessed May 1, 2020

4. Yan Y, Yang Y, Wang F, et al. Clinical characteristics and outcomes of patients with severe covid-19 with diabetes. BMJ Open Diabetes Research and Care 2020;8:e001343.

5. Arentz M, Yim E, Klaff L, et al. Characteristics and outcomes of 21 critically ill patients with COVID-19 in Washington State. Jama 2020;323:1612–4.

6. Osborne TF, Suarez P, Edwards D, Hernandez-Boussard T, Curtin C. Patient Electronic Health Records Score for Preoperative Risk Assessment Before Total Knee Arthroplasty. JBJS Open Access 2020;5:e0061.

7. Osborne TF, Veigulis ZP, Arreola DM, Röösli E, Curtin CM. Automated EHR score to predict COVID-19 outcomes at US Department of Veterans Affairs. Plos One 2020;15.

8. Albandar H, Markert R, Agrawal S. The relationship between aspirin use and mortality in colorectal cancer. Journal of Gastrointestinal Oncology. 2018;9(6):1133–1137.

9. Chow JH, Khanna AK, Kethireddy S, et al. Aspirin Use is Associated with Decreased Mechanical Ventilation, ICU Admission, and In-Hospital Mortality in Hospitalized Patients with COVID-19. Anesthesia & Analgesia. 2020.

10. Wichmann D, Sperhake J-P, Lütgehetmann M, et al. Autopsy findings and venous thromboembolism in patients with COVID-19: a prospective cohort study. Annals of internal medicine 2020.

11. Li Y, Wang M, Zhou Y, et al. Acute cerebrovascular disease following COVID-19: a single center, retrospective, observational study. Stroke Vasc Neurol 2020.

12. Mohamed-Hussein AA, Aly KM, Ibrahim M. Should aspirin be used for prophylaxis of COVID-19 induced coagulopathy? Medical Hypotheses 2020;144:109975-

13. Koupenova M, Kehrel BE, Corkrey HA, Freedman JE. Thrombosis and platelets: an update. European heart journal. 2017 Mar 14;38(11):785–91.

14. Glatthaar_JSaalmüller B, Mair KH, Saalmüller A. Antiviral activity of aspirin against RNA viruses of the respiratory tract—an in vitro study. Influenza and other respiratory viruses 2017;11:85–92.

15. Little P. Non-steroidal anti-inflammatory drugs and covid-19. British Medical Journal Publishing Group; 2020.

16. The use of non-steroidal anti-inflammatory drugs (NSAIDs) in patients with COVID-19: scientific brief, 19 April 2020: World Health Organization; 2020. Report No.: WHO/2019-nCoV/Sci_Brief/NSAIDs/2020.1. Accessed July 24, 2020

17. Pickett KE, Wilkinson RG. Income inequality and health: a causal review. Social Science & Medicine. 2015 Mar 1;128:316–26.

18. Elgar FJ, Stefaniak A, Wohl MJ. The trouble with trust: Time-series analysis of social capital, income inequality, and COVID-19 deaths in 84 countries. Social Science & Medicine. 2020 Oct 1;263:113365.

